# PIE Toolbox: SSM-PCA Based Software for PET Diagnostic Pattern Analysis

**DOI:** 10.64898/2026.05.28.26354341

**Authors:** Mikhail Romanov, Maxim Kireev, Mikhail Didur, Denis Cherednichenko, Alexander Korotkov, Pedro Valdes-Sosa, Qing Fan, Qing Wang, Alzheimer’s Disease Neuroimaging Initiative

## Abstract

One of the prominent methods in neuroimaging data processing is SSM-PCA, which is based on principal component analysis and allows for the identification of diagnostically significant patterns in the form of statistical maps. We developed software, PIE Toolbox, employs SSM-PCA and classification based on the obtained diagnostic patterns revealed from functional and structural tomographic brain imaging. The program supports the entire analysis pipeline including preprocessing of brain images, diagnostic patterns extraction, building classification models, and prediction based on them. The resulting diagnostic patterns are weighted principal components obtained through SSM-PCA, or their linear combinations. PIE Toolbox allows selection of relevant structural and functional brain patterns, computation of their expression values in regions of interest, classification using support vector machines, and evaluation of model performance via cross-validation. This approach enables the use of patterns as features of intergroup differences for individual diagnosis. The software has been validated on both simulated and ADNI datasets.

**One Sentence Summary:** The parcellation-based modification of SSM-PCA outperformed the standard SSM-PCA method in PET image classification, as demonstrated through benchmarking on the ADNI database.

## INTRODUCTION

Structural and functional human brain neuroimaging has made a huge contribution to understanding the mechanisms of brain diseases and their application in clinical practice. In recent years, a many software and hardware solutions have been actively designed and developed. However, various methodological factors often limit their application in real clinical practice for diagnostic purposes.

There are different approaches in hands-on neuroimaging data analysis. Methods that enable the identification of important disease-related factors appear particularly promising. Multivariate approaches excel at this task because they capture complex interdependencies between multiple variables. One of these methods is SSM-PCA (Scaled Subprofile Model and Principal Component Analysis). SSM-PCA makes it possible to obtain components associated with a specific factor from the data (e.g., disease presence). These components appear as statistical spatial maps, known as diagnostic patterns, and can be considered as an indirect characteristic of the inner functional restructuring of the brain *(1, 2)*. SSM-PCA allows to factorizing dataset to a group of patterns that are supposedly reflecting different factors within this dataset, thus potentially providing separation of relevant patterns and non-relevant covariate features. SSM-PCA excels at revealing functional connectivity patterns because it analyzes voxel-wise covariance, capturing network-level alterations across the brain that univariate methods miss.

A key advantage of SSM-PCA patterns over other multivariate methods is their high interpretability. Patterns are three-dimensional voxel images in which the values of the voxels reflect intergroup differences, with positive values indicating a relatively increased signal intensity and negative values indicating a relatively decreased signal intensity in one group versus another. Due to the SSM decomposition, this method allows one to evaluate changes in functional connectivity, rather than absolute changes in the signal *(3)*.

However, SSM-PCA approach has several drawbacks that limits its application for research and diagnostic purposes. In most cases, the expression of the pattern is calculated as the expression throughout the brain for each subject. This means that each pattern corresponds to only one value per subject, reflecting the degree of “similarity” between the subject and the pattern. The main limitation of this approach is that a pattern can show significant differences at the group level, like in univariate analysis, yet there is considerable overlap between individual values. This overlap can hinder reliable classification of intermediate cases *(4)*. This is especially relevant for clinical data where patient data are highly heterogeneous and when dealing with early stage of disease progression. Moreover, SSM-PCA is an unsupervised learning algorithm, and pattern extraction does not require a priori knowledge of the target variable. This means that multiple patterns may correlate with a single feature, while a single pattern may simultaneously combine multiple features (e.g., the presence of a disease) *(5)*.

Thus the classical SSM-PCA often does not accurately reflect the distinguishing features of the revealed pattern and produces only one value per subject. This reduces the effectiveness of the model, particularly in heterogeneous groups where features may be differentially distributed across group patterns and individual patterns confound multiple features. Although SSM-PCA excels at analyzing changes at the group level, these limitations complicate the transition from group-based to individual-based diagnostics.

The expression values in the SSM-PCA are calculated as single scalars per subject, rendering the classification univariate. Thus, each subject is projected onto one dimension, frequently causing overlap between groups. To achieve greater flexibility, a shift to multivariate classification is needed. This involves two steps: first, considering multiple patterns; second, assessing not only entire patterns but also regions of interest to yield multiple variables per subject rather than a single scalar. This approach simultaneously accounts for contributions from multiple patterns as well as interregional connectivity.

A promising approach is to obtain expression values within predefined regions of interest by subdividing a whole-brain statistical pattern into several subregions. In this way, a single global pattern is transformed into multiple functional submaps, which retain the relationships captured by the original pattern but offer greater flexibility in interpretation and analysis. Assessing expression separately within these subregions can provide a more detailed characterization of functional rearrangements and may better reflect individual variability between subjects.

To make full use of this decomposition, a robust classification method is needed. One of the promising approaches is the combination of SSM-PCA with the Support Vector Machine (SVM) classifier. SVM can provide reliable classification and high precision in association with SSM-PCA; it has been reported that SVM outperforms methods such as K-Nearest Neighbors, Random Forest, and Naive Bayes in FDG-PET-based classification *(6–8)*.

Currently, no publicly available SSM-PCA-based solutions provide a universal pipeline that covers diagnostic pattern extraction, analysis, classification model construction, validation, and evaluation. It creates the necessity to develop end-to-end workflow that seamlessly integrates preprocessing, masking, SSM-PCA pattern derivation, classifier training, and rigorous validation.

Currently, there are three main software packages designed for SSM-PCA analysis. The most popular is ScAnVP (Scan Analysis and Visualization Processor), developed by the Feinstein Institutes for Medical Research. ScAnVP is able to construct diagnostic patterns using the SSM-PCA method, export results, calculate the expression of the pattern for new subjects, and analyze them *(9)*.

Another software is MATLAB-based “Generalized Covariance Analysis,” which includes SSM-PCA with features like logarithmic transformation, normalization, and Ordinal Trend Canonical Variance Analysis *(10)*. The SSM-PCA analysis is also included in the RESTplus software for the MATLAB platform (version 1.24 and higher). RESTplus is designed to process fMRI data *(11)*.

A major limitation of the above-mentioned toolboxes is focusing on univariate data analysis, where pattern expression is calculated for the entire brain ignoring regional specificity. Although SSM-PCA is a multivariate method, its whole-brain expression values are univariate scalars per subject and pattern. This limits subsequent analyzes to univariate statistics, despite the availability of multiple pattern scores for richer multivariate modeling. It can limit interpretative power and decrease the efficacy of individual diagnostics. Thus, developing a flexible, accessible tool enabling multivariate SSM-PCA analysis, interpretation, and classification with regional expression focus is essential for enhancing research and clinical applicability of such class of non-univariate approaches.

## RESULTS

### PIE Toolbox

Since there is currently no open-source software that allows full cycle of creating SVM classification models based on the SSM-PCA method and validating our proposed pipeline, we developed our own solution, PIE Toolbox (Pattern Identification and Evaluation Toolbox) using Python. The software implements functions for SSM-PCA per se, along with pattern selection and combination, expression calculation, classification model creation, optimization, and validation. All the main results described below were obtained and processed using the PIE Toolbox. The methods implemented in the PIE Toolbox are described in detail in the Materials and Methods section.

### Generated data model performance

In the first stage of validation, SSM-PCA analysis was performed with procedurally generated 3D images. These were simple geometric shapes with superimposed noise: both the training (n=15) and control groups (n=15) contained 3D cubes; the anomaly in the training group was simulated by a reduced signal in the form of an ellipsoid within the cube. This setup allowed for testing various aspects of SSM-PCA performance. As expected, PIE Toolbox successfully identified a simulated defect in training group and effectively eliminated additive and multiplicative effects when logarithmic image transformation was enabled.

We included additional groups for hold-out validation: training validation group (n=5) and control validation group (n=5). All the resulting patterns matched theoretical expectations, and the classification models built from them demonstrated 100% accuracy.

### ADNI dataset model performance

Procedurally generated images allow for verification of the basic functions of the PIE Toolbox, allowing modeling of predetermined patterns and the influence of additional factors. However, this approach does not assess the applicability of PIE Toolbox to real clinical data and cannot be considered as robust evidence of the method’s viability. To address this issue, we validated the software using data from the open dataset.

For validation, we used data from ADNI subjects with Alzheimer’s disease and healthy volunteers. Inclusion criteria comprised group membership according to ADNI metadata, availability of static PET ^18^F-FDG images co-registered with MRI, and absence of significant imaging artifacts. The main and validation groups were selected to ensure a balanced number of subjects per group.

Two groups of 152 individuals each were formed, from which 10 subjects were randomly selected for validation. Thus, the analysis included AD (n=142) and HC (n=142) groups, with AD-val (n=10) and HC-val (n=10) reserved for hold-out validation. The separation procedure and all subsequent steps were repeated 100 times in Monte Carlo cross-validation.

**Table 1.** Characteristics of subject data.

|  | n | Gender (M/F) | Age |
| --- | --- | --- | --- |
| AD | 142 | 85/57 | $75.37 \pm 8.18$ |
| HC | 142 | 70/72 | $75.03 \pm 6.20$ |
| AD-val | 10 | 6/4 | $73.70 \pm 7.92$ |
| HC-val | 10 | 4/6 | $74.80 \pm 4.87$ |

To prevent data leakage and contamination of the classifier, PET-FDG images from the AD-val and HC-val groups were excluded from both the SSM-PCA analysis and the development of the classification mode. All images were normalized to the MNI space and smoothed with a full width at half maximum (FWHM) of 6.0. Preprocessed images were visually inspected to exclude potential artifacts.

After performing SSM-PCA, the patterns accounting for the first 50% of the explained variance were selected, with a maximum of 20 patterns allowed. Three patterns were found, explaining 19.51%, 16.85%, and 13.38% of variance, respectively. Expression between the AD and HC groups differed significantly only in the first pattern (p < 0.001). Based on the selected patterns, a combined pattern was computed using logistic regression. The resulting pattern structure corresponded to previous findings of SSM-PCA analyses on the ADNI dataset *(12)*.

**Figure 1.**
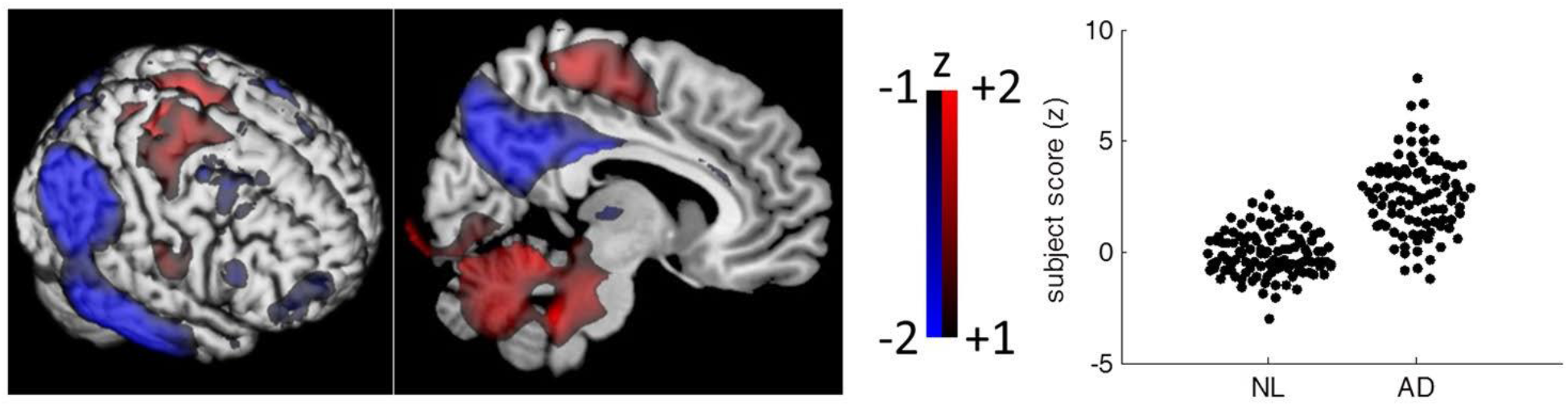
The combined SSM-PCA AD-pattern and its expression scores, printed from Katako et al., «Machine Learning Identified an Alzheimer’s Disease-Related FDG-PET Pattern Which Is Also Expressed in Lewy Body Dementia and Parkinson’s Disease Dementia» *(12)*.

**Figure 2.**
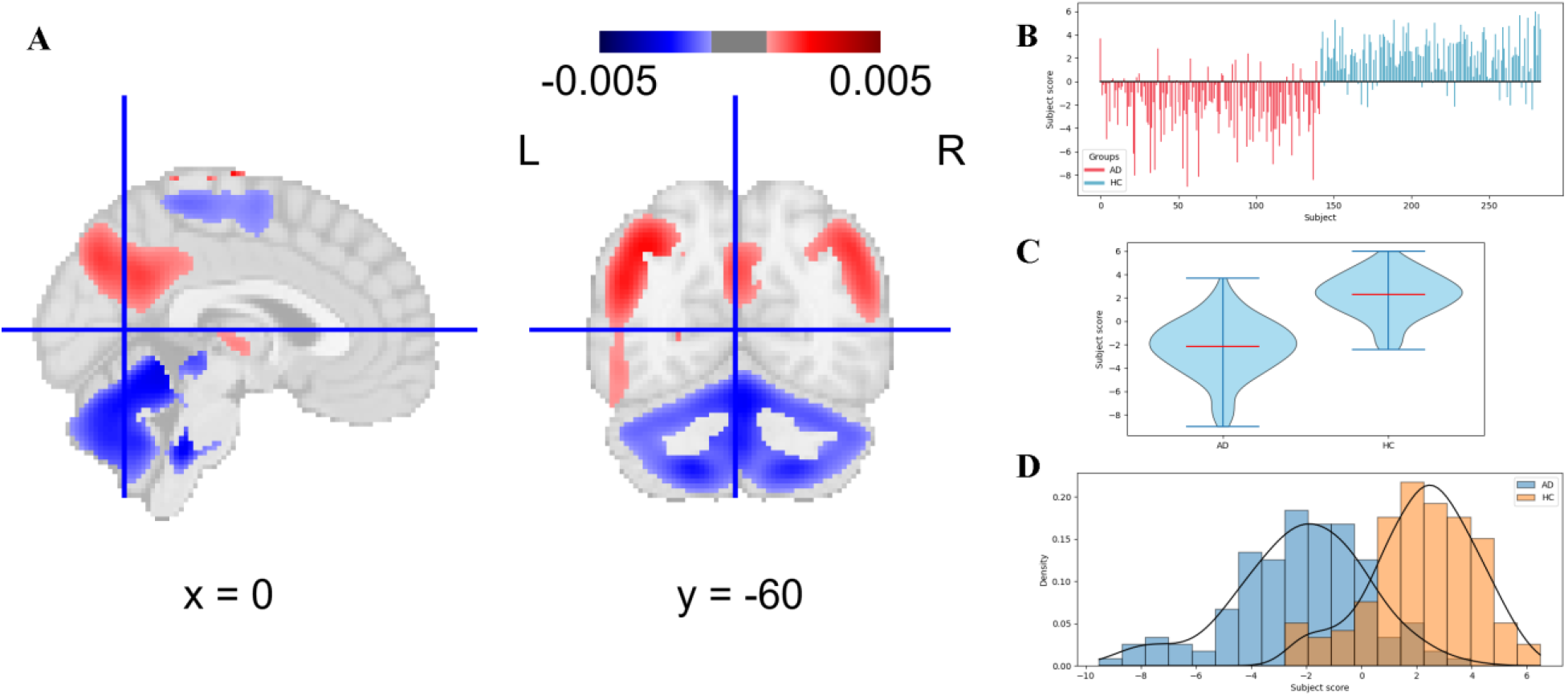
(A) Visual representation of the combined pattern; (B) Bar plot of individual expression scores; (C) Violin plot of expression scores; (D) Histogram of expression scores. The pattern structure and expression value distributions closely resemble those in Figure 1. The scores The standard SSM-PCA method computes whole-brain pattern expression values, equivalent to the sum of single-voxel expressions across the brain. Classification models were constructed using whole brain expression values to validate the approach and assess performance.

After this stage, classification models were built using several parcellation maps: the AAL atlas (Automated Anatomical Labeling) *(13)*, the Brainnetome Atlas *(14)*, and also parcellation map derived from the combined pattern automatic segmentation. Pattern expression was calculated within regions of interest (ROI) derived from these brain atlases. ROIs that did not include any voxels from the combined pattern were excluded from further analysis. Table 2 presents results voxel-wise pattern expression throughout the entire brain, the data-driven parcellation, AAL, and Brainnetome atlases. Metric values were obtained through Leave-One-Out cross-validation and Hold-out validation (using AD-val and HC-val groups).

**Table 2.** Effectiveness of PIE Toolbox classification models. Values presented as Leave-One-Out/Hold-Out validation metrics.

| Performance metrics (Leave-One-Out/Hold-out) | Whole Brain | Data-driven parcellation | AAL | Brainnetome atlas |
| --- | --- | --- | --- | --- |
| Sensitivity | 0.82/0.80 | 0.87/0.90 | 0.89/0.9 | 0.91/1 |
| Specificity | 0.85/0.90 | 0.87/1 | 0.85/1 | 0.89/1 |
| Accuracy | 0.83/0.85 | 0.87/0.95 | 0.87/0.95 | 0.90/1 |
| AUC | 0.86/0.91 | 0.90/1 | 0.92/1 | 0.94/1 |

### Comparison with ScAnVP

In order to validate the patterns obtained using PIE Toolbox, the data from data from procedurally generated 3D images (simple geometric cubes with superimposed noise and simulated anomalies) and the ADNI dataset was analyzed in between group manner.

During comparing the patterns obtained by the PIE toolbox and the ScAnVP software, the Pearson correlation coefficient equaled 1 for all pairs of patterns revealed. This indicated that the structure of the patterns obtained with PIE Toolbox is identical to that of ScAnVP. This perfect correlation indicates identical pattern structures, thereby confirming the validity of the PIE Toolbox in accurately reproducing established SSM-PCA neuroimaging patterns.

### Monte Carlo cross-validation

We conducted Monte Carlo cross-validation with 100 iterations (Table 3). These results were consistent with our previous findings. The leave-one-out and hold-out validation methods showed no significant differences. The model based on whole-brain expression performed the worst compared to other models. Consistent with previous results, predefined atlas-based models proved superior, with the Brainnetome model slightly outperforming the AAL model, although the advantage was negligible. The model based on data-driven parcellation showed no improvement compared to the whole pattern model.

**Table 3.** Mean effectiveness and standard deviation of PIE Toolbox models. Monte Carlo validation with 100 iterations of Leave-One-Out/Hold-Out cross-validation. Values presented as Leave-One-Out/Hold-Out validation metrics with corresponding standard deviations.

| Performance metrics (Leave-One-Out/Hold-out) | Whole Brain | Data-driven parcellation | AAL | Brainnetome atlas |
| --- | --- | --- | --- | --- |
| Sensitivity | 0.88±0.01/<br>0.87±0.10 | 0.87±0.01/<br>0.86±0.01 | 0.89±0.01<br>0.90±0.10 | 0.90±0.01/<br>0.90±0.09 |
| Specificity | 0.87±0.02/<br>0.85±0.11 | 0.87±0.01/<br>0.87±0.01 | 0.89±0.01<br>0.88±0.10 | 0.89±0.01/<br>0.88±0.11 |
| Accuracy | 0.87±0.01/<br>0.86±0.08 | 0.87±0.00/<br>0.87±0.09 | 0.89±0.01<br>0.89±0.07 | 0.89±0.01/<br>0.89±0.07 |
| AUC | 0.89±0.01/<br>0.88±0.10 | 0.89±0.01/<br>0.89±0.09 | 0.94±0.00<br>0.95±0.05 | 0.95±0.00/<br>0.95±0.05 |

### Outlier correction

To assess outlier impact, we compared patterns explaining the first 50% of cumulative variance, along with their combined patterns, with no correction, after 10 Robust PCA iterations, and after 100 Robust PCA iterations. Full cohorts were used (AD, n=152; HC, n=152). Combined patterns followed the standard algorithm for coefficient computation.

Mean voxel-wise differences between each pair of patterns (both individual 50% variance and combined) across no correction, 10 iterations, and 100 iterations did not exceed 0.2% of standard deviations of the original images. No significant differences emerged in efficacy metrics: sensitivity, specificity, and balanced accuracy were identical with and without correction, while AUC varied by no more than 0.07%. These changes were considered negligible, indicating the dataset lacked anomalies requiring correction.

## DISCUSSION

We developed and implemented a software algorithm that combined SSM-PCA with multivariate SVM models. The models were trained on PET FDG data obtained from 304 subjects in the ADNI dataset and demonstrated a high binary healthy/AD classification performance. Moreover, we demonstrated that switching to a multivariate classifier, by extracting expression values within ROIs, significantly improves classification accuracy.

The combination of SSM-PCA and SVM has previously been applied to various neurological diseases, including Parkinsonism *(6)*, Alzheimer’s disease, Lewy body dementia, frontotemporal dementia *(12, 15)*, etc. However, previous studies typically used whole-brain expression values, thus employing univariate classification based on the severity of the patterns. In contrast, our approach employs regional multivariate analysis.

Although multivariate classification for PET images has been explored in previous research by separating independent components into regions of interest, these studies used independent component analysis rather than SSM-PCA *(16)*, which has limitations associated with its assumption of statistical independence of diagnostic patterns. Therefore, potential of SSM-PCA remained underexplored for multivariate classification tasks, and the current methodological study based on PIE Toolbox application addresses this gap by introducing a comprehensive framework with pattern selection, combination and optimized classification modeling.

In the first stage of validation, we verified that the results met expectations under predefined and controlled parameters using procedural data. This confirmed that the core functionality of PIE Toolbox matches the source data parameters and performs as predicted. Nevertheless, procedural data do not allow us to fully assess software performance under real clinical conditions. We tested different variants of the SSM-PCA/SVM models, including ROI-based approaches. We have selected the AAL and Brainnetome atlas for SSM-PCA with ROI extraction. The AAL atlas is probably the most well-known and widely used map for image parcellation. It is integrated into major neuroimaging software packages, including SPM, MRIcron, and the Nilearn Python package. Previous studies have demonstrated the utility of the AAL atlas for feature extraction in classification pipelines *(15, 17)*. Consequently, AAL was selected as a reference standard to ensure the reproducibility and comparability of the results, which is essential for a robust model performance assessment.

The Brainnetome atlas represents a more advanced alternative, as it incorporates structural and functional connectivity information. Thus, it can potentially provide most efficient method for segmenting the patterns. We performed feature extraction with AAL and Brainnetome atlases to evaluate the consistency of the model outcomes.

The results obtained were consistent with those described in previous studies. In line with our expectations, classification based on whole-brain patterns was quite effective but was inferior to classification based on regions of interest. The classifier derived from ROIs extracted from patterns outperformed whole-pattern classification in voxel, but remained inferior to approaches using expression scores from predefined atlases. The classifier based on the Brainnetome atlas was found to be the most effective based on accuracy (89% with Monte-Carlo validation) and AUC (0.95 with Monte-Carlo validation).

We hypothesize that the superiority of predefined atlases over data-driven is explained by several factors. Firstly, heterogeneity may exist within pattern regions with comparable signal changes, making differentiation difficult at the intergroup level. Secondly, voxels with values close to zero are removed from the images during thresholding. It is possible that some areas, despite contributing minimally to the overall pattern expression due to low variance, still reflect important functional rearrangements. Such areas are likely to be excluded when constructing a parcellation map from the pattern but retained when using a predefined atlas. It is important to note that the optimal pattern partitioning approach may vary depending on the specific model used.

Using the ADNI data set allowed us to compare our results with those described in previous studies. The studies used different sets of performance metrics, so we based our comparisons on available data. In the study by Katako et al., SSM-PCA analysis was performed for ADNI FDG PET data. For the first SSM-PCA pattern obtained, classification sensitivity and specificity were 0.755 and 0.829, respectively, with an AUC of 0.849; the combined pattern improved these metrics to 0.830 sensitivity, 0.847 specificity and 0.897 AUC. Additionally, a voxel-wise SVM classifier was performed employing ISDA (Iterative Single Data Algorithm) and sequential minimal optimization (SMO) was conducted. SVM-ISDA showed a sensitivity of 0.840 and a specificity of 0.955, while SVM-SMO achieved 0.894 and 0.892, with AUCs of 0.945 and 0.939, respectively, which is closely match the PIE Toolbox Brainnetome atlas-based classification *(12)*.

In the work by Illán et al., a modification of PCA was used to obtain patterns and the SVM model from ADNI data. Unlike SSM-PCA, the first step was to calculate the mean image vector in each group, which was then processed using PCA. Thus, the analysis included group-average images rather than individual subject images. Expression values were obtained in a manner similar to that of SSM-PCA. The accuracy of classification for Alzheimer’s disease versus healthy volunteers using SVM with an RBF kernel was 88.24%, with sensitivity and specificity of 88.64% and 87.80%, respectively, which is compatible with the PIE Toolbox whole-brain pipeline but inferior to Brainnetome atlas results *(18)*.

Other approaches based solely on SVM classification with ADNI datasets demonstrated variable results: Cabral et al. achieved accuracy of 66.78% with ternary SVM classification *(19)* while Dukart et al. employed SVM for binary classification, reporting an accuracy of 87.5% based on predefined regions of interest *(20)*. These studies illustrate the versatility of SVM in handling both multi-class and binary classification tasks in Alzheimer’s disease research, with lower accuracy varying depending on the dataset and classification scheme.

Thus, the model we obtained based on the whole voxel-wise pattern was comparable in efficiency to the SSM-PCA models described in the literature, with the accuracy of the PIE Toolbox 0.83 to0.85 and 0.67 to0.88 in previous studies. The classifier that was performed best based on the Brainnetome atlas, according to cross-validation and hold-out validation, was more effective than the classification models considered based on SSM-PCA and SVM, achieving an accuracy of 0.90-1 and an AUC of 0.94-1.

Obtained results demonstrate that calculating expression in regions of interest may improve the quality of classification compared to calculating expression across the entire brain. We extract data-driven parcellation map as the regions of pattern with similar weights with thresholding and combining neighbor voxels using hyperparameter tuning. However, the resulting map was outperformed by the AAL and Brainnetome atlases.

Possible explanations for this effect could be as follows. The thresholding could exclude voxels with small absolute weights that are important to the final model. Moreover, because voxels with similar weights tend to be treated as one unified group, problems can arise when there are subtle variations within this region. For example, the cerebellum can be treated as one huge area of hypermetabolism, which is true for group-wise comparison, but the anatomic atlas treats it as a cluster of smaller areas and thus reflects changes of the functional network inside the cerebellum which is more precise.

In contrast, using a predefined atlas for parcellating of the patterns may be suboptimal without prior knowledge of relevant ROIs. Although pattern-based maps can miss low-weight areas and merge regions with similar weights, they better capture the intrinsic structure of the pattern. Therefore, we recommend selecting a predefined atlas based on the results of the pattern-based approach. Pattern-generated ROIs help identify the most suitable atlas for the final model.

SSM-PCA is a well-established method for classification and feature extraction, offering considerable flexibility. Its patterns have been validated for various conditions and diseases; however, it has seen limited clinical application due to complex initial setup, lack of user-friendly open-source interfaces, and challenges with heterogeneous real-world data. Our PIE toolbox addresses these limitations and surpasses previous approaches by integrating SSM-PCA with multivariate SVM classification. The PIE toolbox offers an SSM-PCA pipeline enhanced by ROI-specific pattern expression for multivariate classification, experimental Monte Carlo validation and outlier correction features for researchers, and design optimized for potential future integration into clinical workflows.

### Limitations and Future Research

SSM-PCA is a robust and highly interpretable method, and ROI-based expression calculation can greatly enhance its effectiveness. However, there are some limitations that may hinder the application of the PIE Toolbox pipeline to clinical practice.

First, SSM-PCA patterns are generally consistent across different cohorts, but models calibrated on data acquired with a specific scanner may be less effective when applied to data from another scanner. This issue can be addressed in two ways. The first is to run the PIE Toolbox pipeline on global multicenter data; however, the resulting models are highly likely to reflect only the most common and prominent features, potentially overlooking traits specific to individual scanners.

Therefore, global models should be considered robust but imprecise tools. The second approach is to derive a “hardware norm”, i.e. to calculate models specific to individual scanners. Hardware norms are expected to be more precise, but must be calculated separately for each scanner. They are also more likely to give poorly trained models due to potential data scarcity. Based on the foregoing, we propose that the optimal algorithm should combine both methods: estimating expression using thoroughly validated global patterns and correcting hardware bias with local patterns.

Secondly, PCA is sensitive to outliers. Thus, even a single outlier image can distort all diagnostic patterns. During the prediction stage, an outlier image may be interpreted as a highly abnormal sample and misclassified. This issue is particularly relevant for neurodegenerative disorders, which are associated with brain atrophy and can produce images with inconsistent signal decreases. In this study, we performed robust PCA *(21)* and determined its impact to be negligible compared to the non-robust variant. However, in datasets exhibiting prominent outliers, such correction may be necessary. For less consistent data, multiple tests are recommended to evaluate the method performance. Future validation across diverse imaging datasets, with varying numbers of iterations, is essential to ensure the deployment of robust PCA within the default PIE toolbox pipeline.

Parameters such as the percentage of explained variance and statistical significance between groups are often used to evaluate the effectiveness of patterns. We believe that these metrics, while helping the researcher notice the most relevant patterns, may inefficiently capture important intergroup characteristics. Explained variance reflects total dataset variability and blind to groups, allowing non-target factors to dominate, while statistical tests perform well for groups but, being group-focused, cannot be applied to individual subjects.

Calculating expression at the within-subject level can, in turn, help to conduct the intergroup analysis stage more efficiently. Using the weights obtained from the classification and regression models, it is possible to interpret the patterns and highlight the regions most important for classification, even if they exhibit relatively small variability between subjects. This is especially relevant for heterogeneous groups, where small differences within groups can be lost amid intersubject variability. Furthermore, the proposed system can serve as a basis for analyzing and predicting not only qualitative but also quantitative data through regression models.

## MATERIALS AND METHODS

### ADNI database

This research required FDG-PET data that has been rigorously validated through prior studies and are freely available in open access. It was essential to estimate the robustness of the PIE Toolbox pattern extraction and its concordance with previous results.

Data used in the preparation of this article were obtained from the Alzheimer’s Disease Neuroimaging Initiative (ADNI) database (adni.loni.usc.edu). The ADNI was launched in 2003 as a public-private partnership led by Principal Investigator Michael W. Weiner, MD. The primary goal of ADNI has been to test whether serial magnetic resonance imaging (MRI), positron emission tomography (PET), other biological markers, and clinical and neuropsychological evaluation can be combined to measure the progression of mild cognitive impairment (MCI) and early Alzheimer’s disease (AD).

ADNI is a repository of neuroimaging and clinical data that includes PET images. The inclusion criteria for the present study were the availability of baseline FDG-PET images of sufficient quality from patients diagnosed with Alzheimer’s disease or healthy controls. The groups were composed of equal numbers of subjects from ADNI 1, 2, and 3 datasets.

### Preprocessing

Images were preproced by ADNI, resulting in coregistered scans with intensity normalization, uniform resolution, and consistent voxel sizes; no smoothing was applied.

The images for SSM-PCA must be spatially normalized into a common coordinate space, typically the Talairach or MNI space (Montreal Neurological Institute), and smoothed *(9)*. We performed spatial image normalization int the MNI 152 space using the Advanced Normalization Tools (ANTs) module. We used the SyN method, which applies both the affine and deformable transformations of the image to match a template. After registration, smoothing with a Gaussian kernel (Full Width at Half Maximum 6.0) was performed, also using the ANTs module.

### Image masking

To perform SSM-PCA analysis, voxels that do not contain useful data must be excluded using a mask. The PIE Toolbox supports three methods for creating and applying binary masks: calculating a mask from a threshold on the original data, using a predefined binary or probabilistic mask, or using a mask derived from a brain atlas.

For threshold mask creation, a binary mask is first generated for each image based on a specified relative threshold (from 0 to 100 percent, where 100% is the maximum absolute value within the image). A separate mask was created for each image. In the second step, all masks are multiplicatively combined: if a voxel is excluded in at least one mask, it is excluded from the final mask.

The PIE Toolbox supports mask definition in two ways: by loading a predefined binary or probabilistic mask, or by constructing a mask from a neuroimaging atlas through the selection of specific regions of interest. In the probabilistic case, the mask is binarized using a chosen relative threshold.

These methods can be used simultaneously, with the final mask being the voxel-by-voxel product of the resulting binary masks. Each image undergoes this masking procedure, and the resulting set of voxels is used directly for SSM-PCA. This approach aligned with the standard SSM-PCA preprocessing, ensuring that only relevant voxels are included in the analysis.

### SSM-PCA

SSM-PCA is a modification of PCA (Principal Component Analysis) used to extract diagnostic patterns from a set of three-dimensional images. PCA normalizes the data and extracts an individual profile for each image, eliminating additive and multiplicative global variations at both the group and the subject levels. Based on these individual profiles, a statistical map is constructed that reflects changes in functional connectivity.

The SSM Model (Scale Subprofile Model) was described by Moeller et al., that was used to decompose a set of 3D images into a GMP (group average profile), an SRPs (individual residual profile) and a scalar value of GSFs (Global Scaling Factor) *(22)*:

P_sv_ = GSF_s_*(GMP_v_+SRP_sv_)

where P is an s×v matrix (s is the total number of subjects, v is the number of voxels in one 3D image after applying the mask).

In this case, the 3D image must be represented as a vector of length v, where each element of the vector represents a voxel value. Voxels irrelevant to the analysis are excluded using a binary mask as described above.

Further transformation was performed using the PCA method. PCA extracts the principal components, which are linear combinations of the original features ordered by decreasing variance *(9)*. Using the PCA method, SRP can be considered as the product of GIS vectors (group invariant subprofiles, which in SSM-PCA correspond to the principal components) and scalar values of Subprofile Scaling Factor(SSF) *(22)*. Each pattern is formed by multiplying a principal component by the square root of its singular value.

To calculate principal components, SSM-PCA uses subject-to-subject covariance matrix with dimensionality s×s where s is a number of subject *(5, 23)*. As a result, the number of patterns obtained with SSM-PCA is equal to the number of subjects although it may be limited to a smaller number.

Thus, SSM-PCA consists of the following steps *(9, 22, 24)*:

1. Using a mask to exclude voxels from the analysis, as described above;
2. Transforming each subject’s 3D image into a vector of length v. Form a matrix P of size s×v, where each row represents one subject’s masked voxels;
3. Logarithmic transformation of the data (this step can be skipped if the original data donot require logarithmic transformation): P_sv_ → Log P_sv_
4. Data are grouped by rows subtracting the mean of each row from its values, producing a matrix centered on the row.. The data is then represented as a deviation from the row mean Q_sv_ = Log P_sv_ - mean_v_(Log P_sv_) where Q is a row-centered matrix;
5. Data column centering is performed similarly to row centering. A column initially represents the set of values for a given voxel for each subject after logarithmic transformation and row centering. SRP_sv_ = Q_sv_ - GMP_v_ where GMP_v_ = mean_s_(Log Q_sv_).
6. Decomposing the resulting SRP matrix using PCA into GIS vectors (principal components) and Scalar Subprofile Scaling Factors (SSF);
7. The pattern is the product of the corresponding GIS vector and the square root of its singular value. Although the GIS and its corresponding SSFs can be used directly as the pattern and expression values, respectively, this step enables comparison of different patterns by taking their explained variance into account. This ensures that patterns are weighted according to their contribution to the overall data structure, making it possible to assess their relative importance in the analysis;
8. The expression of a pattern for a particular subject is the scalar product of the SRP and the corresponding pattern.

GIS vectors are represented in Z-score format, where a value of zero represents the average weight of the voxels through subjects and regions. The GIS vector reflects the deviation from the mean rather than the absolute values *(22)*. Scalar SSF values can be used to examine differences between the control and patient groups for each GIS using standard hypothesis testing methods.

If the the difference between groupsis significant, GIS can be interpreted as a characteristic pattern of one of the groups, and SSF as the expression of that pattern in each subject *(9)*. According to Eidelberg et al., the SSF obtained for each subject is strongly correlated with the severity of the disease *(3)*. The positive voxel weights in the obtained pattern indicate a positive correlation between the expression value of the subject and the corresponding signal, while the negative weights indicate a negative correlation of the pattern expression *(25)*.

As stated previously, the SSF represents the whole expression of the pattern for subjects included in the analysis. For clinical use, this expression must also be calculated for new subjects. This is done by applying logarithmic transformation (if necessary) and normalizing the new data by the GMP vector obtained previously during SSM-PCA. The projection of the new subject’s data onto the pattern vector then gives its expression value.

The standard SSM-PCA algorithm calculates expression across the entire pattern but can also be performed for individual regions. The general expression is equal to the sum of the expression values of each individual voxel. This means that the sum of scores across all regions is equal to the whole-brain score as long as no voxels are excluded from the calculation. Therefore, it is possible to divide the pattern into multiple ROIs and assess scores region-wise. However, this raises the issue of selecting relevant ROIs.

### Extracting regions of interest

Regions of interest can be obtained using a predefined atlas. However, this method has a significant drawback: the regions of interest may not coincide with the regions where relevant structural rearrangements are observed. Since choosing an atlas suitable for a specific study can be a non-trivial task, PIE Toolbox allows to build a data-driven parcellation map directly from the selected patterns.

This process consists of three main stages. In the first stage, all voxels whose absolute value is less than a certain threshold (specified as a relative or absolute value) are removed from the pattern. In the second stage, adjacent voxels in which the difference is less than the specified parameter are combined into a region of interest. In the third stage, regions of interest with a small number of voxels are excluded. The connected-components-3d module is used for the second and third stages *(26)*.

The PIE Toolbox allows for setting parameters for voxel exclusion, merging, and region-of-interest selection. Since optimal values may vary between different studies, they should be selected individually for each case. The PIE Toolbox includes tools for automatic parameter selection (see the section “Parameter optimization”).

### Support vector machine

The support vector machine (SVM) is a supervised learning algorithm that classifies data by constructing a separate hyperplane *(27)*. This method demonstrates high accuracy in the automated classification of neurodegenerative diseases from ^18^F-FDG PET data (78% and 80% for pattern-based and ROI-based approaches, respectively, in Perovnik et al. *(15)*, 53–82% across ROIs in Yang et al. *(28)*, 73% in Booth et al. *(29)*, high accuracy is also reported in some meta-analyses *(20, 30)*) and in the rat study by Devrome et al. *(31)*, making it the method of choice for PIE Toolbox. Using SVM, it is possible to build a model for classifying neuroimaging data across multiple variables.

The current version uses the method implemented in the scikit-learn 0.24.0 module. The module allows using SVM to classify two or more groups by customizing the kernel and regularization parameter C. For classification, the toolbox uses the support vector machine (SVM) (C-Support Vector Classification) with a hard margin (C=1.0) and the RBF or linear kernel. The variables used are the average values within the regions of interest obtained in the previous steps. The efficiency of the obtained model can be assessed using both cross-validation and Hold-out Validation.

Cross-validation in the PIE Toolbox is performed using the Leave-One-Out and StratifiedKFold methods. Cross-validation simulates the presence of a test set to evaluate the performance of the model. When using Leave-One-Out validation, only one image is included in the test set. This method produces more accurate results, but is more resource-intensive. When using the StratifiedKFold method, the set is split into a selected number of parts (5 by default), and each part is used as a test set in turn. Each test set contains representatives of each class in approximately the same proportion as the full set. The metrics obtained during cross-validation are the average values of the metrics from the models obtained or their derivatives *(32)*. We used StratifiedKFold only as a preliminary method due to its low accuracy and did not utilize it for the final results.

Since the original images are used to derive the pattern, cross-validation methods cannot ensure that images from the test set do not contaminate the resulting models. Validation on a hold-out sample is carried out using PIE Toolbox prediction tools. The program predicts hold-out images and compares the results with the reference labels, after which it calculates the classification performance. This is the most reliable method as it avoids data leakage.

The method evaluates the membership score of each subject in a specific class. Furthermore, a cross-validation is performed which calculates the probability of each subject’s membership in each group. It is should be noted that the group obtained by the classifier may not coincide with the most probable group for the subject. In this case, the result should be considered insufficiently reliable, which means that the subject cannot be confidently classified into one group or another.

### Parameter optimization

Variables used for classification in PIE Toolbox represent the average expression values of patterns in the specified regions of interest. However, the challenge of selecting relevant regions of interest that match the structure of the patterns obtained is a complicated task.

The PIE Toolbox includes an iterative algorithm to optimize the parameters. Each iteration is an exhaustive search on a given grid. For each parameter, a set of values within a given interval is determined, after which enumeration is caried out through all possible combinations of parameter values. For each combination, a classification model is built and cross-validated. The combination with the best result (balanced accuracy is calculated by default) and the fewest variables is considered the optimal combination for that iteration.

Parameters can be specified in three ways:

- Fixed value: The parameter is set to a single value and remains unchanged throughout the iterations.
- A set of parameter values: The algorithm iterates over a predefined set of values, which are constant throughout the iterations.
- Parameter interval. A sequence of uniformly distributed values within a specified interval, with intervals becoming narrower between iterations.

At each subsequent iteration, the grid of parameters specified as an interval is updated. The interval size is calculated as:

S_i +1_ = S_i_ * k

where S is the interval size, i is the number of the last completed iteration, k is the interval narrowing coefficient (default 0.7).

The minimum and maximum values of the new interval are calculated, equal to

Val_min_ = Best_i_ – 0.5*S_i +1_

Val_max_ = Best_i_ + 0.5* S_i +1_

where is Val_min_ and Val_max_ — new minimum and maximum values of the interval, Best_i_ is the optimal value of the parameter at the last completed iteration.

If Val_min(i +1)_ is less than the value Val_min (1)_ specified for the first iteration, Val_min(i +1)_ is set to the value Val_min(1)_. The same happens if Val_max(i +1)_ exceeds the initial maximum value.

Thus, at each iteration, a new, narrower interval is constructed, centered on the optimal parameter value from previous iterations. This adaptive approach, compared to a standard grid search, reduces the number of parameter combinations to be tested while maintaining high accuracy. With a single iteration, the optimization tool behaves identically to a simple grid search.

After selecting patterns, it is necessary to set parameters that determine the relative threshold, the delta value, and the minimum voxel count in the region of interest. The PIE Toolbox employs the parameter optimization procedure described above to identify the optimal combination for classification. Before starting the analysis, the intervals within which the parameters will vary, as well as the number of divisions for these intervals, must be specified. Based on the analysis results, optimal ranges for the threshold and delta can be determined for optimal classification.

The most effective model is selected for the subsequent stages of analysis.

### Model effectiveness

The PIE Toolbox allows to save confusion matrices, reflecting the correspondence between the classification during cross-validation and the true labels. The matrices are saved in a file in .xlsx format as a table; in addition, the performance values are calculated for each group: sensitivity, specificity, precision, balanced accuracy, positive predictive value (PPV), negative predictive value (NPV), and area under the curve (AUC).

Accuracy reflects the overall probability of correctly classifying a subject and is calculated as (TP + TN) / (TP + TN + FP + FN), where cases classified into a given group are denoted as follows: TP — true positives, TN — true negatives, FP — false positives, and FN — false negatives *(33)*.

Sensitivity is the probability of correctly classifying a positive result and is calculated as TP / (TP + FN). Specificity is the probability of correctly classifying a negative result and is calculated as TN / (TN + FP) *(33)*.

To eliminate the influence of sample size imbalance, the balanced accuracy is additionally estimated, which is calculated as the arithmetic mean between sensitivity and specificity *(34)*.

In addition, the efficiency is estimated based on the ROC curve (receiver operating characteristic curve) of the model, which shows the relationship between sensitivity and specificity at all possible thresholds. The ROC curve allows one to calculate the AUC parameter. The AUC can be interpreted as a diagnostic information value, with AUC<0.5 corresponding to no diagnostic effectiveness, AUC 0.5−0.7 to weak effectiveness, AUC 0.7−0.8 to moderate effectiveness, AUC 0.8−0.9 to good effectiveness, and AUC 0.9−0.1 to excellent effectiveness *(33, 35)*.

PPV is calculated as the ratio of true positives to all positives: TP / (TP + FP). NPV is calculated as the ratio of true positives to all positives: TN / (TN + FN) *(33)*.

The above metrics are maintained by PIE Toolbox for the researcher to evaluate the effectiveness of the model. The cross-validation process for the resulting classifier is automatically performed using the LeaveOneOut method (by default) or StratifiedKFold, described above. However, a significant drawback of cross-validation based on training data is the risk of data leakage. If the same data used to generate diagnostic patterns is used for training, the patterns may contain indirect information about the subjects being validated. Thus, cross-validation allows one to evaluate the effectiveness of the resulting model, but an objective assessment requires the selection of a special validation group whose data are not included in the SSM-PCA and classifier generation stages. For this purpose, PIE Toolbox supports validation on hold-out data. The evaluation methods and metrics are similar to those described for validation on the training data.

During the group definition stage, we identified the validation groups. This method provides lower accuracy than Leave-One-Out, but it eliminates data leakage. Using the results obtained by both methods, we can evaluate the effectiveness of the classifier with great reliability.

### Monte Carlo cross-validation

Although Leave-One-Out and Hold-Out methods validate classifiers on a given sample, the hold-out sample may differ dramatically from the training sample. This issue is especially relevant for small groups, where the risk of obtaining a non-representative hold-out sample is high. The PIE Toolbox includes an integrated Monte Carlo machine for cross-validation to provide a robust estimate of model consistency.

The integrated Monte Carlo cross-validation (MCCV) pipeline repeats each step of the basic PIE Toolbox pipeline, from generating masks for the image dataset to Hold-Out validation. In every MCCV iteration, the training and hold-out image sets are randomly selected based on a preset percentage or number of images. We used the default option for hold-out subsets, in which the number of images in each validation group is equal, and every group is represented in the hold-out sample. The final result is calculated as the mean of all results in iterations.

MCCV is not suitable for routine research or clinical practice due to its high computational cost. We employ MCCV as a technique to assess the consistency and reliability of the metrics obtained.

### Supported data formats

The PIE Toolbox works with 3D images in NIFTI-1 format. Images must be converted to a single coordinate space (e.g., MNI space, Montreal Neurological Institute). This format imports subject images, masks, and parcellation maps. Diagnostic patterns and the parcellation maps derived from them can be exported in the NIFTI-1 format. To save intermediate results, the PIE Toolbox employs the pickle format, which enables serialization of Python objects. Patterns, parcellation maps, and classification models are exported as pickle files.

The PIE Toolbox includes a set of tools for visualizing and analyzing patterns. A pattern or a parcellation map based on it can be displayed in an HTML page as an interactive statistical map using the nilearn.plotting Python module with a selected threshold value *(36)*. By default, the “seismic” palette is used to display diagnostic patterns, while parcellation maps use a discrete palette calculated programmatically based on the number of retrieved regions.

The expression values of the whole brain pattern can be represented as a bar or violin plot, as well as a double histogram. The bar histogram can be sorted by increasing expression values between subjects. The results of the classification model validation can be exported in .xlsx format as a table.

### Procedural image generation

The test images for validation are simple geometric shapes with 3D noise superimposed on them. For this purpose, a cube with a side length of 20 and a voxel value of 1 is constructed in a 3D array of 100×100×100 elements. For the train group, an ellipsoid with axes ranging from 8 to 12 and a voxel value of 0.6 is also constructed within the cube. Noise is simulated by adding a random value between 0.0 and 0.1 to each voxel value.

In the images obtained in this way, the anomaly is simulated by an ellipsoid with reduced voxel values. The first pattern obtained in SSM-PCA, if the algorithm is successful, should highlight this region as a characteristic change. The procedural generation script is included in the PIE Toolbox.

### The PIE Toolbox and the ScAnVP comparison

To analyze the correlation of patterns obtained using PIE Toolbox and ScAnVP, the patterns were created using identical sets of source data, and a mask was calculated based on a specified threshold to ensure that the masking algorithm was identical. Using a custom Python script, the resulting patterns were converted into one-dimensional vectors, and the Pearson correlation coefficient was calculated between the patterns from PIE Toolbox and ScAnVP. A Pearson correlation coefficient of 1 indicates the complete equality and stability of the implementation of SSM-PCA .in the PIE Toolbox.

### Outlier correction

SSM-PCA relies on principal component analysis (PCA), which is sensitive to outliers and thus reduces model reliability. PCA maximizes variance, and outliers can severely distort results.

The Robust PCA method addresses this by decomposing the input matrix into a low-rank matrix (clean signal) and a sparse matrix (noise and outliers). This method iteratively refines these matrices to minimize reconstruction error, with the final low-rank matrix then processed via standard PCA *(21)*. The described algorithm was implemented as an experimental feature in the PIE Toolbox.

## Data Availability

The data supporting the findings of this study are derived from the Alzheimer's Disease Neuroimaging Initiative (ADNI). ADNI data are publicly available to approved researchers through the Image and Data Archive (IDA).

https://adni.loni.usc.edu/

## Acknowledgments

Data collection and sharing for this project was funded by the Alzheimer’s Disease Neuroimaging Initiative (ADNI) (National Institutes of Health Grant U01 AG024904) and DOD ADNI (Department of Defense award number W81XWH-12-2-0012). ADNI is funded by the National Institute on Aging, the National Institute of Biomedical Imaging and Bioengineering, and through generous contributions from the following: AbbVie, Alzheimer’s Association; Alzheimer’s Drug Discovery Foundation; Araclon Biotech; BioClinica, Inc.; Biogen; Bristol-Myers Squibb Company; CereSpir, Inc.; Cogstate; Eisai Inc.; Elan Pharmaceuticals, Inc.; Eli Lilly and Company; EuroImmun; F. Hoffmann-La Roche Ltd and its affiliated company Genentech, Inc.; Fujirebio; GE Healthcare; IXICO Ltd.; Janssen Alzheimer Immunotherapy Research & Development, LLC.; Johnson & Johnson Pharmaceutical Research & Development LLC.; Lumosity; Lundbeck; Merck & Co., Inc.; Meso Scale Diagnostics, LLC.; NeuroRx Research; Neurotrack Technologies; Novartis Pharmaceuticals Corporation; Pfizer Inc.; Piramal Imaging; Servier; Takeda Pharmaceutical Company; and Transition Therapeutics. The Canadian Institutes of Health Research is providing funds to support ADNI clinical sites in Canada. Private sector contributions are facilitated by the Foundation for the National Institutes of Health (www.fnih.org). The grantee organization is the Northern California Institute for Research and Education, and the study is coordinated by the Alzheimer’s Therapeutic Research Institute at the University of Southern California. ADNI data are disseminated by the Laboratory for Neuro Imaging at the University of Southern California.

AI was used as a secondary tool for final text proofreading.

## Data and materials availability

The PIE Toolbox, including its code and graphical user interface, is openly accessible via the official repository at https://github.com/IHB-IBR-department/PIE_toolbox, which includes procedural validation image dataset. The ADNI dataset employed for benchmarking is publicly available at https://adni.loni.usc.edu/.

